# Analysis of cell-mediated immunity in people with long COVID

**DOI:** 10.1101/2021.06.09.21258553

**Authors:** Nerea Montes, Èlia Domènech, Sílvia Guerrero, Bárbara Oliván-Blázquez, Rosa Magallón-Botaya

## Abstract

**Introduction:** The objective of this study is to analyse the specific immune response against SARS-CoV-2 in those affected by Long Covid (LC), attributable to T cells (cell-mediated immunity) and to carry out a parallel analysis of the humoral response and lymphocyte typing.

**Methodology:** Descriptive cross-sectional study of 74 patients with LC for at least 4 months since diagnosis. The collected data were: information on the COVID-19 episode and the persistent symptoms, medical history and a specific cell-mediated immunity to SARS-CoV-2 through flow cytometry, assessing the release of interferon-gamma (IFN-Ɣ) by T4 lymphocytes, T8 lymphocytes and NK cells. Descriptive and comparative analyses were carried out.

**Results:** Patients with LC had negative serology for Covid-19 in 89% of cases but 96% showed specific cellular immunity to SARS-CoV-2 an average of 9.5 months after infection: 89% of this response corresponded to T8 lymphocytes, 58% to NK cells, and 51% to T4 lymphocyte (20% negligibly positive). Most of them had altered immune cell typing and we found that T4 lymphocyte counts were low in 34% of cases and NK cell high in 64%. Macrophage populations were detected in the peripheral blood of 7% of them. Patients displayed a higher percentage of illnesses related to &[Prime]abnormal&[Prime] immune responses, either preceding SARS-CoV-2 infection (43%) or following it in 23% of cases.

**Conclusion:** The immune system appears to have an important involvement in the development of LC and viral persistence could be the cause or consequence of it. Further analysis with a control group should be performed.

## INTRODUCTION

The SARS-CoV-2 (COVID-19) infection, which has placed the entire world into an unprecedented pandemic situation, continues to be studied, with new findings being made daily as to its different levels of affectation and evolution in different patients.

It is estimated that approximately 10-20% of those affected, after suffering the initial infection, which is often mild or moderate with no need for hospitalisation, continue to show symptoms beyond the expected recovery time for the illness (2-4 weeks). This is in spite of no viral load being detected in the upper respiratory tract through a nasopharyngeal PCR, even over a year after the episode which triggered the symptoms ^(1-3)^.

This condition has come to be known as Long COVID (LC). LC can be defined as the syndrome or condition with which patients that have been infected with SARS-CoV-2 present where, after its acute phase, they continue to show persistent or cyclical symptoms over time that have a significant impact on their quality of life. A significant deterioration of previously healthy and active people’s health is observed, and these patients’ lives become worryingly limited. Its characterisation and clinical management are not yet completely defined ^(3)^. Carrying out research and providing care for this large group of people who have had COVID-19 but continue to live with debilitating symptoms is a social responsibility that must be fulfilled as soon as possible.

LC mainly affects middle-aged people, the majority of whom are female, with multi-systemic affectation including 36 symptoms on average, with up to 200 having been described ^(4,5)^. These include chest pain, palpitations, tachycardia, breathlessness, muscle and joint pain, headaches, cognitive impairment (‘brain fog’), neuropathy, paraesthesia and fatigue, among many others ^(6-9)^. Patients with LC tend to experience a combination of all of these symptoms ^(6,10)^, and it is not known how long they will persist. After the outbreak of severe acute respiratory syndrome (SARS), similar long-lasting symptoms were described (chronic fatigue, pain, weakness, depression and sleep disturbance) ^(11)^.

In spite of this illness being recognised by the WHO as a part of COVID-19, the mechanisms involved in this kind of progression are unknown. Hypotheses have been advanced about viral persistence thanks to a kind of reservoir, or possible alteration of the immune system ^(12)^.

Moreover, a significant number of LC patients who contracted the virus during the first wave (March 2020) did not undergo diagnostic tests, as they did not have access to a PCR within the recommended timeframe. They also often do not have positive serology for the virus (humoral response), which would allow this process to be attributed to an initial SARS-CoV-2 infection ^(13)^.

This suggests a possible alteration of the immune response in this group/sub-group which could hinder effective viral clearance and contribute to the persistence of symptoms over a period of months ^(12)^.

It was therefore decided to study another facet of the specific immune response against SARS-CoV-2 in those affected by LC, attributable to T cells (cell-mediated immunity). This is less widely used, but is playing an increasingly important role ^(14)^ that could confirm prior exposure to SARS-CoV-2 in the LC population that did not have access to diagnostic testing, as well as identifying differential traits that will help us to understand the illness’ aetiopathogenic mechanism.

It was also decided to carry out a parallel analysis of the humoral response and lymphocyte typing in an attempt to complete the immunity profile or to identify the impact on/progression in defence cells after several months of persistent symptoms.

It was considered of interest to explore a possible relationship between this data and the possible aetiological cause of viral persistence in LC patients and the existence of immune-related diseases, either pre-dating or diagnosed following COVID-19. We also wanted to investigate the potential reactivation of latent viruses ^(12)^ and record the symptoms reported at the time of the study, with a view to identifying these patients and the mechanisms involved.

All this could contribute towards improving our understanding of the illness of LC, possibly leading to the discovery of the hypothetical cause of this long-term decline in health.

## METHODOLOGY

### Design

This is a descriptive cross-sectional study of patients with a COVID-19 diagnosis and symptoms that have persisted for at least 4 months since diagnosis.

An online survey designed by the LC Patients of Spain Collective’s scientific-health group was conducted. This was distributed by email to the patients belonging to this collective. Data was collected from February to May 2021.

### Sample and inclusion/exclusion criteria

In total there were 74 participants, drawn from the Long COVID Patients of Spain Collective, who met the following inclusion criteria: a diagnosis of COVID-19 through positive tests (PCR, antigens, serology) or a clinical picture compatible with the disease, followed by multi-systemic and fluctuating symptoms, considered to be Long COVID, for at least 4 months since diagnosis or suspected initial infection. The clinical inclusion criteria have also taken into account that during March and early April 2020, due to a shortage, diagnostic tests were not carried out on a significant number of people with symptoms attributable to COVID-19, and many patients did not require hospitalisation, and therefore did not have access to these tests at that time.

The exclusion criteria were the following: patients experiencing symptoms from 1 to 3 months after the initial infection. This was in order to better define persistent symptoms, distinguishing this from a potential recovery situation if they lasted for only one month, given that the severity of the initial infection has not been quantified.

Duplicate or incomplete survey responses were removed. No other exclusion criteria were considered in order to conduct a study as close to the clinical reality of the LC patient population as possible.

### Study variables

Sociodemographic data (sex and age), and information on the COVID-19 episode and the persistent symptoms experienced were collected, as well as a medical history. All these variables were obtained through the questionnaire. In addition, results of the study of specific cell-mediated and humoral immunity against SARS-CoV-2 were included.

As regards the COVID-19 episode, chronological information was collected on the onset of the clinical picture and the kind of diagnostic test used. The date of analysis sample extraction for the present immunological study was also recorded, between January and early May 2021.

In terms of persistent symptoms, participants were asked to indicate three predominant symptoms, including: tiredness/asthenia; shortness of breath and/or coughing; chest and/or back pain; muscle or joint pain; digestive disorders, urinary disorders, headache/migraines; difficulty concentrating, memory lapses or brain fog; palpitations and/or dizziness/POTS (postural orthostatic tachycardia syndrome); tremors, tingling or weak limbs; skin lesions and/or itchy skin or tongue; vision or eye impairment (itchiness, dry eyes, etc); oto-rhino-laryngological impairment (mouth, ears, nose and throat), change in the senses of taste and/or smell, endocrine disorders (menstruation, diabetes, thyroid, etc), vascular disorders (varicose veins, blood clots). Symptoms consistent with viral persistence or reactivation were also included: presence of fever, frequent chills or shivering, adenopathy (swollen lymph nodes in the neck and armpits), or reactivation of Herpesvirus (zoster and simplex type 1).

The medical history information collected was as follows: previous diagnosis of asthma, allergies, auto-immune diseases, chronic fatigue syndrome (CFS)/myalgic encephalomyelitis (ME)/fibromyalgia, postural tachycardia syndrome (POTS), viral infections such as mononucleosis or symptomatic CMV (Cytomegalovirus) or EBV (Epstein-Barr Virus) infection. New diagnoses of these conditions following infection with SARS-CoV-2 (COVID-19) were also gathered.

With respect to the results obtained from the peripheral blood counts conducted in an approved research laboratory, a study of specific cell-mediated immunity to SARS-CoV-2 was carried out through flow cytometry. This involved exposing a concentrate of lymphoid cells to a protein extract of the virus (M, N and S proteins), assessing the release of interferon-gamma (IFN-Ɣ) by T4 lymphocytes, T8 lymphocytes and NK cells.

#### This study included

- Lymphocyte population typing, recording their percentage value as high, normal or low according to laboratory reference values: CD3 T lymphocytes (10.8 to 34.8%), CD4+ or T4 lymphocytes (51.4 to 77.8%), CD8+ or T8 lymphocytes (15.4 to 38.2%), CD 56 or NK (Natural Killer) cells (9.5 a 20.5%), CD19 and CD14 (lymphocytes and monocytes with combined values of 5.2 to 10.3%). The presence of macrophage populations detected in some patients’ samples was also indicated.
- Cellular response measured through the production of IFN-gamma by T4 and T8 lymphocytes and NK cells against viral proteins M (membrane), N (nucleocapsid) and S (spike), with the specific cell-mediated response considered to be positive if it was over 1%, negligibly significant if it was between 1-1.5%, and significantly positive if it was over 1.5%.
- The humoral response or antibody titre to SARS-CoV-2 was also determined through chemiluminescence EIA (enzyme immunoassay), recorded on the questionnaire as positive or negative, with the reference index value being less than or equal to 0.9 for IgM and less than or equal to 1.5 for IgG.

### Statistical analysis

Firstly, a percentage-based descriptive and comparative analysis was conducted on the data obtained through the form, whereby the different variables were analysed, looking for associations between them and respecting the classification developed based on a set of criteria established to differentiate the immunity profile, based on the results obtained through the immunity study. The tables and figures shown show the cases in absolute values and percentages. For comparative analysis of the different variables collected, the chi-squared statistic or Fisher’s exact test was used (where the theoretical frequencies included frequencies below 5, or where the marginal sums of the data set were very unequal). The comparative analysis conducted was as follows: comparison of cell typing and response in patients with positive and negative diagnostic tests; comparison of T4 lymphocyte typing (normal or low) versus NK cells (normal or high); comparison of T4 response with respect to their typing (normal or low), comparison according to immunity profile groups: medical history and symptom variables.

The statistical analysis was performed using SPSS V.21. and *p*-values < 0.05 were considered statistically significant.

## RESULTS

Of the 74 participants, 86.5% are women, almost half (43%) are between 41 and 50 years old, 26% between 51 and 60, and 24% between 21 and 40 (93% between 21 and 60). The opposite extremes of the age range (under 20 and over 60) each accounted for around 3% of participants.

Symptoms lasting for over 9 months were reported by 82% of patients, and over 7 months in 94.6% of cases. Only 5.4% reported symptoms lasting between 4 and 6 months, and the mean length of symptoms since their onset is 9.5 months.

Most cases, therefore, correspond to the pandemic’s first wave of infection, with no **diagnostic test** available in 76% of cases (56 patients). It was found that 81% of survey respondents declared that they had not had a prior positive **microbiological test** (PCR or viral antigen test) due to a lack of access at that time, despite presenting compatible symptoms and/or having come into contact with individuals infected with COVID-19. In cases where a positive PCR or antigen test was obtained (19%), the mean time since this was performed was 7.9 months (12% took place over 9 months ago, and the remaining 7% between 1 and 9 months ago).

In addition, 81% of respondents reported that they did not have a positive **antibody test** either, either because they had not been tested, or because they had tested negative. Cases of positive serology (19%) date back an average of 4.7 months, with 11% of the total number of patients surveyed reporting between 1 and 6 months, 2.7% less than one month ago and the remaining 5.4% more than 7 months ago. Of the 24% that had some kind of prior positive test (18 of the 74 cases), 5 only had a positive PCR or antigen test, 10 patients also had positive serology and 3 only had a positive antibody titre.

The predominant **persistent symptoms** (participants were requested to indicate only 3) among participants were asthenia (66%), muscle or joint pain (47%), palpitations and/or dizziness (42%), brain fog (41%), migraines (31%), dyspnoea and/or coughing (28%) and digestive disorders (23%), followed by chest or back pain or pressure (18%).

Symptoms with lower instances included tremors/tingling or weak limbs (14%), eye disorders (14%), vascular disorders (11%), oto-rhino-laryngological disorders (9%) and endocrine disorders (7%), followed by urinary or skin-related symptoms on 4% and anosmia/ageusia on 3%.

Furthermore, symptoms compatible with viral persistence or reactivation were recorded, finding that 64% of patients suffered from fever, frequent chills or shivering, and around 31% from adenopathy and the reactivation of Herpesvirus (zoster and Herpes simplex type 1).

The figure below refers to **diseases related to ‘abnormal’ immune system responses** (asthma, allergies and autoimmune diseases), either pre-existing or diagnosed after contracting COVID-19 (**Figure 1**).

**Figure 1:**
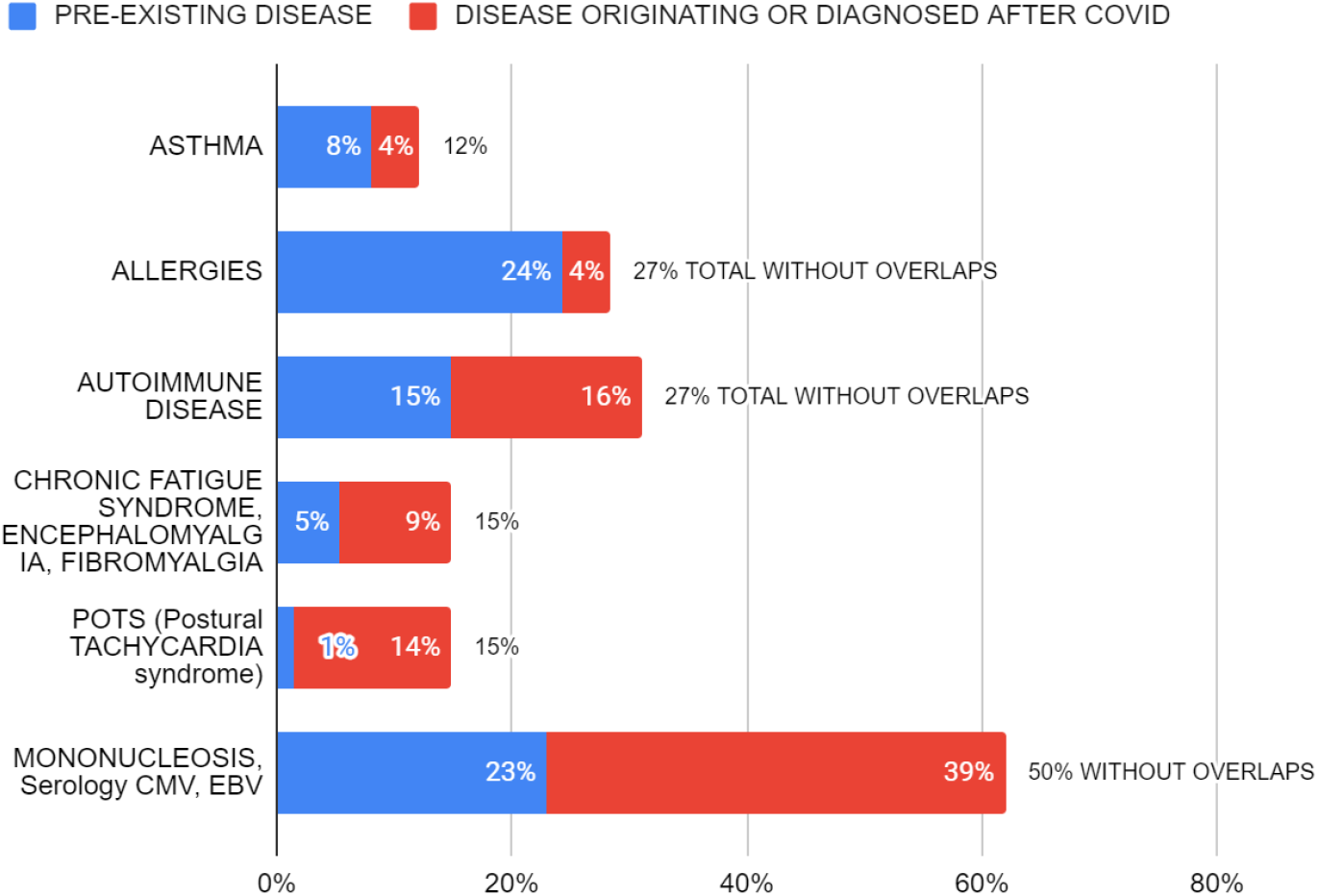
PRE-EXISTING DISEASES VERSUS THOSE ORIGINATING OR DIAGNOSED AFTER COVID-19.

In terms of allergies, 24% of cases were pre-existing. This was true of 15% of autoimmune diseases and, to a lesser extent, 8% of asthma cases. Cases of asthma, allergies, and autoimmunity account for **43% of the total (32/74)**, since 3 patients suffer from both asthma and allergies.

After contracting COVID-19, autoimmune disease has been diagnosed in 16% of patients surveyed, and asthma or allergies in 4% of them. The total number of cases of these diseases after infection with SARS-CoV-2 accounts for **23% of the total sample (17/74)**, as 1 patient suffers from both allergies and autoimmune disease.

In figure 1, the incidence of these diseases is further analysed by adding together their prevalence figures prior to and after infection with COVID-19 (counting overlapping cases only once). The figures obtained are 12% for asthma, and 27% for allergies and autoimmune diseases (totals without overlapping cases on the right-hand side of the graph).

Diagnosis prior to and after infection with SARS-CoV-2 of diseases such as **chronic fatigue syndrome (CFS)** and **postural tachycardia syndrome (POTS)** was also recorded. More cases of CFS were diagnosed after contracting COVID-19 (9%) than beforehand (5%). In the case of POTS, incidence was higher after infection, at 14% compared to 1% beforehand.

The possible relationship between long COVID and the **reactivation of latent Herpesviruses** (Cytomegalovirus/CMV or Epstein-Barr Virus/EBV) was analysed, identifying previous symptomatic episodes of mononucleosis in 23% of patients, and current positive serology against CMV or EBV in 39%, including 2.7% for IgM, 31% for specific IgG and 5.4% for both IgM and IgG against the same viruses. Adding together the prevalence of these latent or reactivated viruses among respondents, this constitutes 50% of the total group, counting those who had prior symptomatic infection and have current positive serology only once, since in 12% of the patients both conditions coincide.

Results were obtained from the **analytical immunity study** conducted from January to early May 2021, with the following findings:

In terms of the **<u>humoral response</u>**, patients with persistent symptoms, in the current analysis, had negative serology for SARS-CoV-2 in 89% of cases (Figure 2.2). However, 8 respondents (11%) had positive serology, with 6 of them having specific IgM an average of 9.7 months after the initial COVID-19 episode (only IgM in 2 of these cases corresponding to 2.7% and combined with IgGs in the 4 other cases corresponding to 5.4%), as well as 2 (2.7%) with only positive IgG (outlined in table 1, where reference is made to the cell-mediated immunity group corresponding to the subsequent classification).

**Table 1:** LOSS OF +ANTIBODIES BETWEEN PREVIOUS TEST AND CELL-MEDIATED IMMUNITY TEST

**SPECIFIC LONG COVID STUDY WITH POSITIVE ANTIBODIES IN THE CELL-MEDIATED RESPONSE TEST**
| Age | Time in months between onset of symptoms and Cellular test with Positive Antibodies | Group NK, T4, T8 | Group NK, T4, T8 | Pre-existing Positive Antibodies | IgM+ post | IgG+ post |
| --- | --- | --- | --- | --- | --- | --- |
| 41-50 | Over 9 months | NK+,T4+,T8+ | Group 1 | +ve pre-existing AB for 1-3 months | IgM+ |  |
| 51-60 | Over 9 months | NK+,T4+,T8+ | Group 1 | +ve pre-existing AB for less than 1 month | IgM+ | IgG+ |
| 51-60 | Over 9 months | NK+,T4-,T8- | Group 4 | +ve pre-existing AB for 4-6 months | IgM+ | IgG+ |
| 51-60 | Over 9 months | NK-,T4+,T8+ | Group 5 | +ve pre-existing AB for 4-6 months | IgM+ |  |
| 31-40 | Over 9 months | NK-,T4+,T8+ | Group 5 |  | IgM+ | IgG+ |
| 41-50 | From 7-9 months | NK-,T4+,T8+ | Group 5 | +ve pre-existing AB for less than 1 month | IgM+ | IgG+ |
| 51-60 | Over 9 months | NK-,T4+,T8+ | Group 5 | +ve pre-existing AB for 1-3 months |  | IgG+ |
| 61-70 | Over 9 months | NK-,T4+,T8+ | Group 5 |  |  | IgG+ |
| 51.3 | 9.7 months for IgM;<br>9.8 months for IgM+IgG |  |  |  |  |  |
| Mean Age | Mean time between onset of symptoms and Cell Response Test |  |  |  |  |  |

**LONG COVID WITH PRE-EXISTING +VE ANTIBODIES WHICH DO NOT REGISTER IN THE CELL RESPONSE TEST**
| Age | Time in months between onset of symptoms and Cellular test with Positive Antibodies | Group NK, T4, T8 | Group NK, T4, T8 | Pre-existing Positive Antibodies | IgM+ post | IgG+ post |
| --- | --- | --- | --- | --- | --- | --- |
| 31-40 | From 4-6 months | NK+,T4-,T8+ | Group 3 | +ve AB 4-6 months ago |  |  |
| 51-60 | Over 9 months | NK+,T4-,T8+ | Group 3 | +ve AB 7-9 months ago |  |  |
| 41-50 | Over 9 months | NK+,T4-,T8+ | Group 3 | +ve AB 1-3 months ago |  |  |
| 41-50 | Over 9 months | NK-,T4+,T8+ | Group 5 | +ve AB 1-3 months ago |  |  |
| 41-50 | Over 9 months | NK-,T4+,T8+ | Group 5 | +ve AB 4-6 months ago |  |  |
| 41-50 | Over 9 months | NK-,T4-,T8+ | Group 7 | +ve AB 7-9 months ago |  |  |
| 41-50 | Over 9 months | NK-,T4-,T8- | Group 8 | +ve AB 7-9 months ago |  |  |

Antibodies lost in 5.4 months on average

A relationship was also found between the results of this serology and that obtained in previous diagnostic tests, with 6 of the 8 (75%) already having tested positive for antibodies in a previous test conducted less than 1 month ago in 2 cases, between 1-3 months ago in another 2, and between 4-6 months ago in the remaining 2 (table 1).

However, apart from these 6 patients with previous positive serology who currently retain this, another 7 had positive antibody titres prior to the current study, and have now lost these, within an average of 5.4 months.

In reference to the cell-mediated immunity study analytical results, figures 2.5 to 2.10 show the typing and specific responses of the different lymphocyte populations.

**Figure 2.1.**
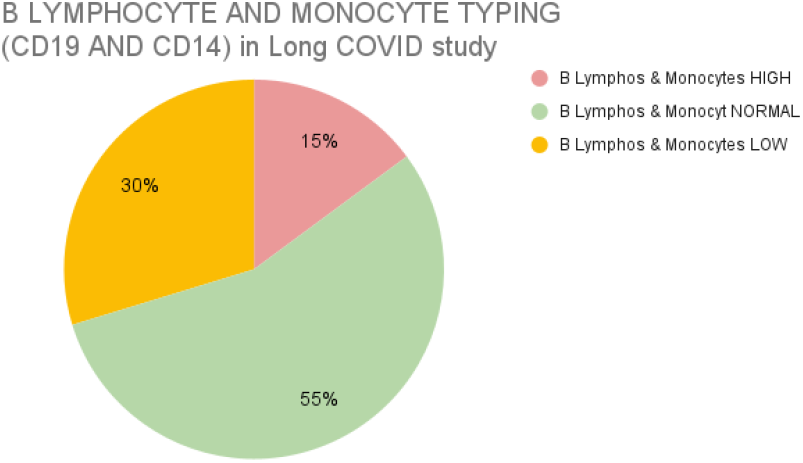
B Lymphocytes and monocytes.

**Figure 2.2.**
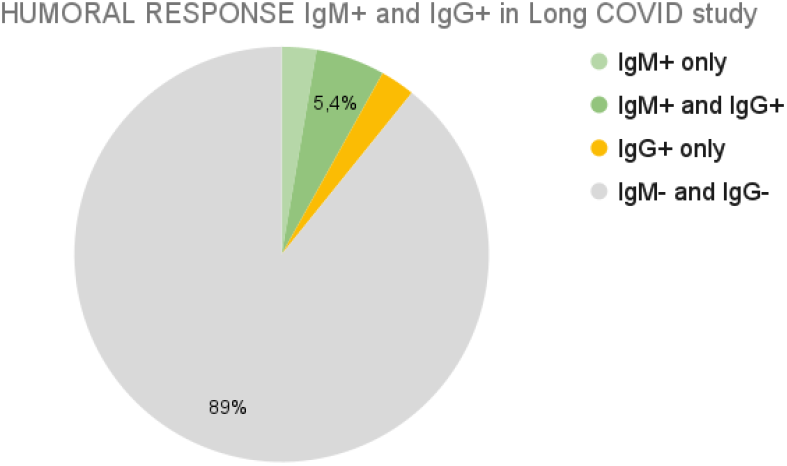
Humoral response.

### As regards the <u>immune cell typing</u> analysis

B lymphocytes and monocytes (Figure 2.1) are normal in 55% of cases, low in 30% and high in 15% of participants.

T lymphocytes (CD3) (Figure 2.3) are normal in 57% of cases, high in 39% and low in 4%. An altered CD3+ T lymphocyte count was found in 43% of participants.

T4 lymphocyte counts (Figure 2.5) show values that are mainly within the normal range (66%), although they are low in 34% of cases, and high values were not found in any participant.

T8 lymphocytes (Figure 2.7) show mostly normal levels; they are high in 26% of cases, and low in the minimum percentage of cases (2.7%), according to the limits established by this laboratory (<15.4%). This means that almost 30% of the patients surveyed have a CD8+ T population that is either abnormally high or low, with the vast majority being abnormally high.

The NK cell count (Figure 2.9) is high in 64% of LC patients and normal in the rest of them. No low count was found.

None of the patients had a high T4 lymphocyte count or a low NK cell count, and significant differences are not observed (p-value = 0.567) when comparing T4 lymphocyte typing (normal or low) with NK cells (normal or high).

In addition, macrophage populations (Figure 2.4) are detected in the peripheral blood of 7% of patients analysed, when they are naturally found in body tissues.

**Figure 2.3.**
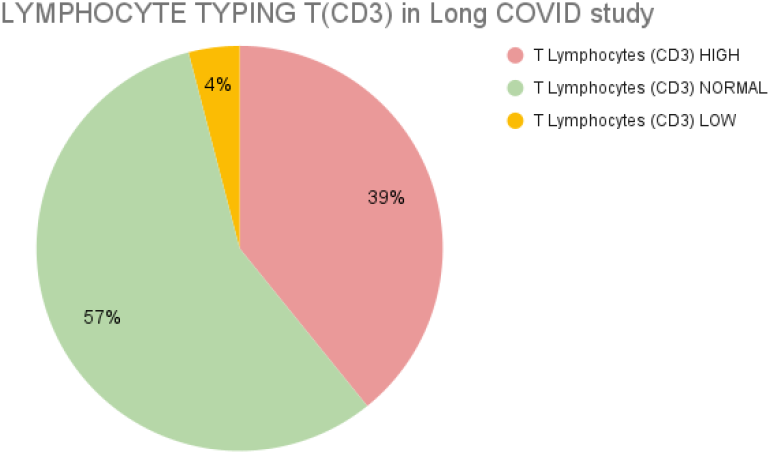
T Lymphocytes (CD3)

**Figure 2.4.**
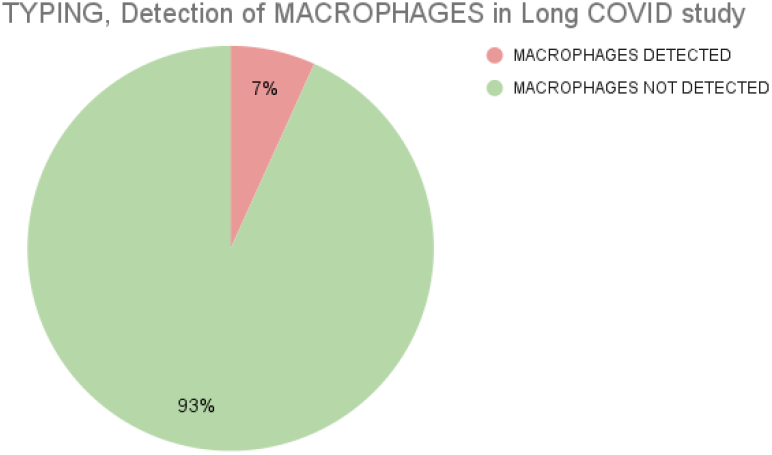
Macrophages.

**Figure 2.5.**
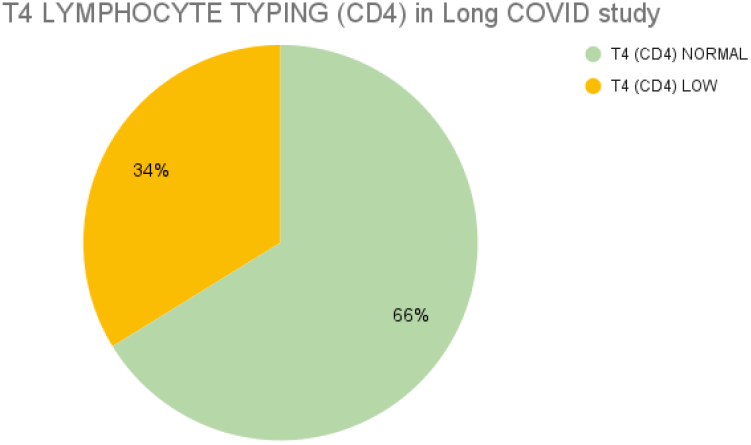
T4 Lymphocytes (CD4): mainly normal; low in a considerable percentage (34%). **Not high in any case**.

**Figure 2.6.**
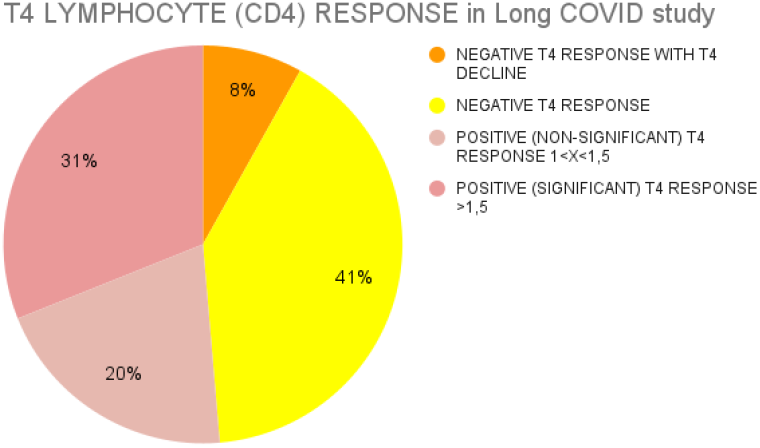
T4-specific response: Negative in almost half of the patients (49%), also showing a decline in populations after exposure to the viral antigen in 8% of cases. Positive in 51% of cases (less than 1.5% response in 20% of them).

**Figure 2.7.**
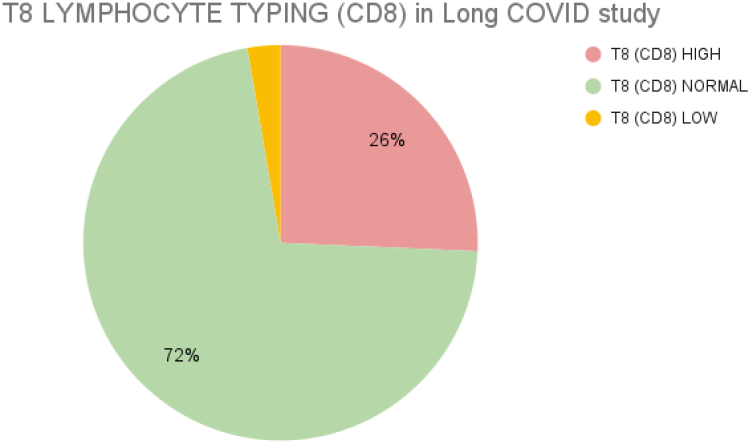
T8 Lymphocytes (CD8): mainly normal, high in 26% of cases and low in 2%, according to the limits established by this laboratory (<15.4%).

**Figure 2.8.**
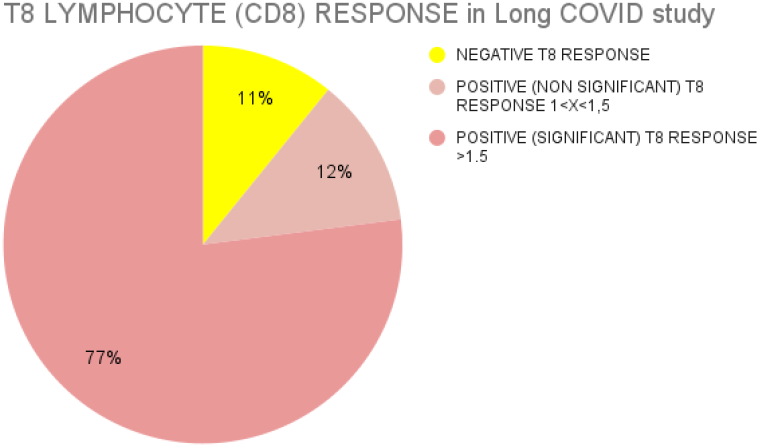
T8-specific response: Positive in 89% of patients (over 1.5% response in 77% of them).

**Figure 2.9.**
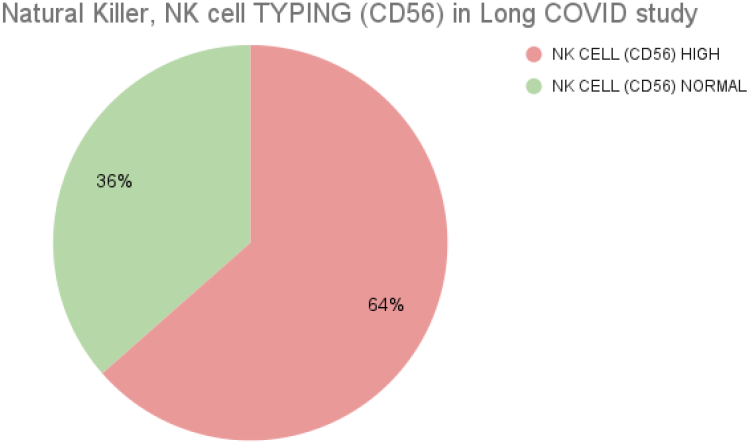
NK Cells: **High in 64%** of patients and normal in the rest. **Not low in any case.**

**Figure 2.10.**
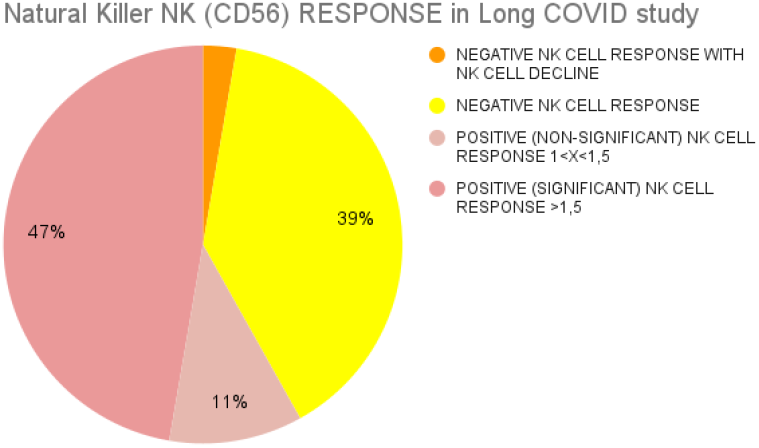
NK Response: **Positive in 58% of cases**, and negative in the remaining 42%. A decrease in cell populations after exposure to the virus was observed in 3% of cases.

As regards cell-mediated immunity, there is a **<u>specific cell-mediated response</u>** against SARS-CoV-2 for T4 lymphocytes (Figure 2.6) positive in 51% of participants, at a negligibly significant level (<1.5% response) in 20% of the total group. As for the negative response (49%), a small number of patients were identified (6/74: 8%) in which T4 lymphocyte populations decrease in number when exposed to the viral antigen.

Likewise, it was statistically proven that there is no significant difference (p-value = 0.266) in the response of CD4+ T lymphocytes among those who have a low or normal count.

In the case of T8 (Figure 2.8), there was a positive result in 89% of cases, significantly so in 77% of them (>1.5% response). As for the NK response (Figure 2.10) this was positive in 58% of patients, and significant in the majority of those cases (47% of total patients). In 2 cases where there was a negative response (2.7%), a decrease in NK populations was observed upon exposure to viral proteins.

Statistical tests confirmed that there are not significant differences in terms of the typing and kind of specific cell-mediated response between the group that had a positive diagnostic test from the time of infection or through subsequent serology, and those who did not undergo a test during the first wave due to lack of access, or who tested negative (table 2). The statistical tests were applied for this purpose, obtaining a p-value > 0.05 for all parameters.

**Table 2:** Typing and Cell-mediated Response by no. and % of cases, confirming that not having a previous +ve test does not influence the results.

| T Lymphocytes (CD3) HIGH | T Lymphocytes (CD3) NORMAL | T Lymphocytes (CD3) LOW | T4 Lymphocytes (CD4) HIGH (N/A) | T4 Lymphocytes (CD4) NORMAL | T4 Lymphocytes (CD4) LOW | T8 Lymphocytes (CD8) HIGH | T8 Lymphocytes (CD8) NORMAL | T8 Lymphocytes (CD8) LOW | NK Cell (CD56) HIGH | NK Cell (CD56) NORMAL | NK Cell (CD56) LOW (N/A) | B Lymphocytes and Monocytes HIGH | B Lymphocytes and Monocytes NORMAL | B Lymphocytes and Monocytes LOW | PRESENCE OF MACROPHAGES | NEGATIVE T4 RESP. WITH T4 DECLINE | NEGATIVE T4 RESPONSE. | POSITIVE (Non-significant) T4 RESP. 1<X<1.5 | POSITIVE (Significant) T4 RESPONSE >1.5 | NEGATIVE T8 RESPONSE | POSITIVE (Non-significant) T8 RESP. 1<X<1.5 | POSITIVE (Significant) T8 RESPONSE >1.5 | NEGATIVE NK RESP. WITH NK DECREASE | NEGATIVE NK RESPONSE | POSITIVE (Non-significant) NK RESP. 1<X<1.5 | POSITIVE (Significant) NK RESPONSE >1.5 | IgM+ HUMORAL RESPONSE to COVID | IgG+ Had Humoral Response to COVID |  |
| --- | --- | --- | --- | --- | --- | --- | --- | --- | --- | --- | --- | --- | --- | --- | --- | --- | --- | --- | --- | --- | --- | --- | --- | --- | --- | --- | --- | --- | --- |
| 39% | 57% | 4% | 0% | 66% | 34% | 26% | 72% | 3% | 64% | 36% | 0% | 15% | 55% | 30% | 7% | 8% | 41% | 20% | 31% | 11% | 12% | 77% | 3% | 39% | 11% | 47% | 8% | 8% | TOTAL |
| 5 | 11 | 2 | 0 | 13 | 5 | 4 | 14 | 0 | 13 | 5 | 0 | 4 | 8 | 6 | 2 | 2 | 5 | 4 | 7 | 3 | 2 | 13 | 1 | 8 | 1 | 8 | 5 | 5 | 18 cases WITH Pre-Existing Positive Test |
| 24 | 31 | 1 | 0 | 36 | 20 | 15 | 39 | 2 | 34 | 22 | 0 | 7 | 33 | 16 | 3 | 4 | 25 | 11 | 16 | 5 | 7 | 44 | 1 | 21 | 7 | 27 | 1 | 1 | 56 cases WITHOUT Pre-existing Positive Test |
| 29 | 42 | 3 | 0 | 49 | 25 | 19 | 53 | 2 | 47 | 27 | 0 | 11 | 41 | 22 | 5 | 6 | 30 | 15 | 23 | 8 | 9 | 57 | 2 | 29 | 8 | 35 | 6 | 6 | 74 cases in TOTAL |
| 28% | 61% | 11% | 0% | 72% | 28% | 22% | 78% | 0% | 72% | 28% | 0% | 22% | 44% | 33% | 11% | 11% | 28% | 22% | 39% | 17% | 11% | 72% | 6% | 44% | 6% | 44% | 28% | 28% | 18 cases WITH Pre-Existing Positive Test |
| 43% | 55% | 2% | 0% | 64% | 36% | 27% | 70% | 4% | 61% | 39% | 0% | 13% | 59% | 29% | 5% | 7% | 45% | 20% | 29% | 9% | 13% | 79% | 2% | 38% | 13% | 48% | 2% | 2% | 56 cases WITHOUT Pre-existing Positive Test |
| 39% | 57% | 4% | 0% | 66% | 34% | 26% | 72% | 3% | 64% | 36% | 0% | 15% | 55% | 30% | 7% | 8% | 41% | 20% | 31% | 11% | 12% | 77% | 3% | 39% | 11% | 47% | 8% | 8% | 74 cases in TOTAL |

Based on the study participants’ cell-mediated immune response to SARS-CoV-2, a classification was made which is set out in the tree diagram in table 3. This begins with the positive or negative specific cell-mediated NK response, or innate immunity (where negative includes reduction of lymphocytes), with a subsequent sub-division according to the CD4+ T lymphocyte response, and these in turn according to the CD8+ T response. A total of **8 differentiated groups** are hereby identified (7 in **figure 3, no patient in group 6**). Likewise, the group to which those who had antibodies and those in whom macrophage populations were detected in peripheral blood belong is specified in the table.

**FIGURE 3:**
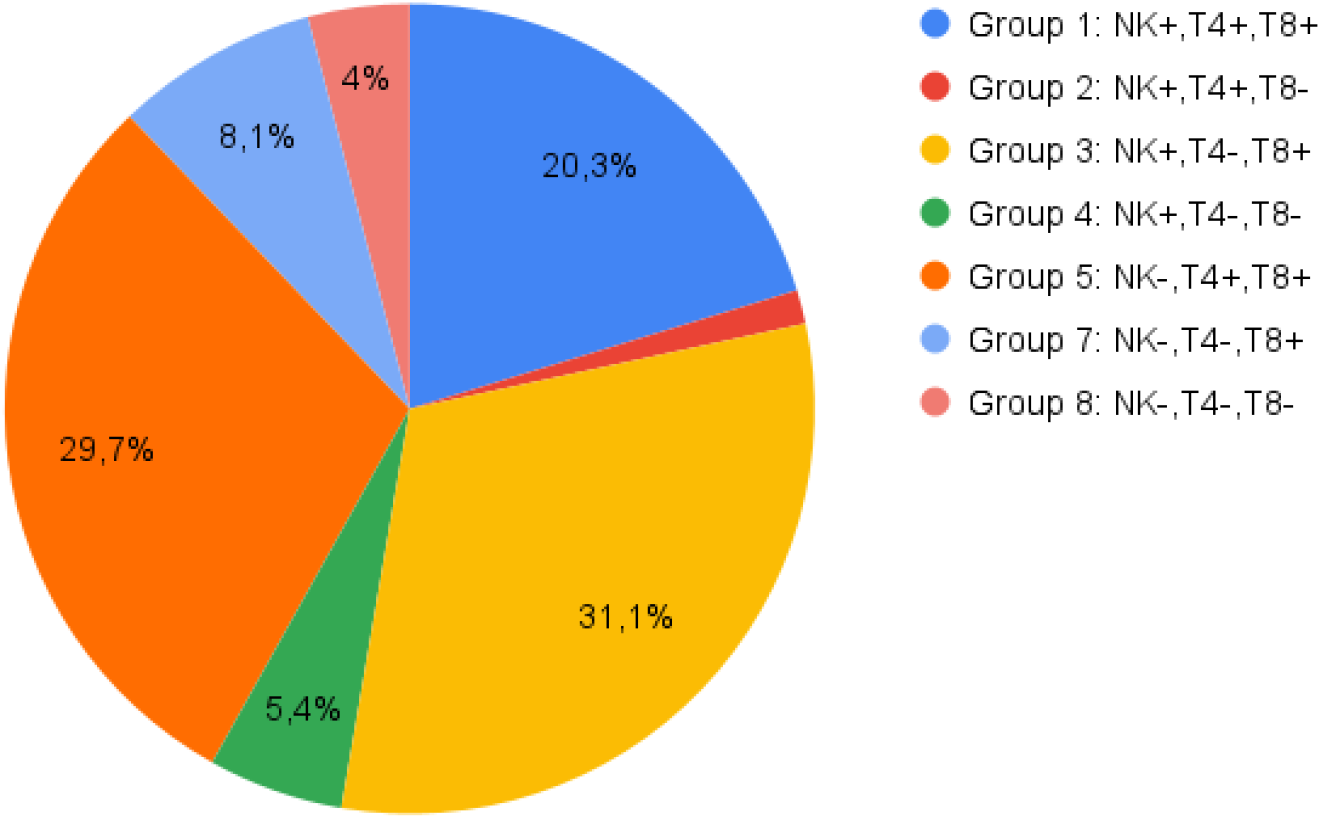
CLASSIFICATION OF GROUPS ACCORDING TO NK IMMUNE PROFILE → T4 → T8.

**TABLE 3:**
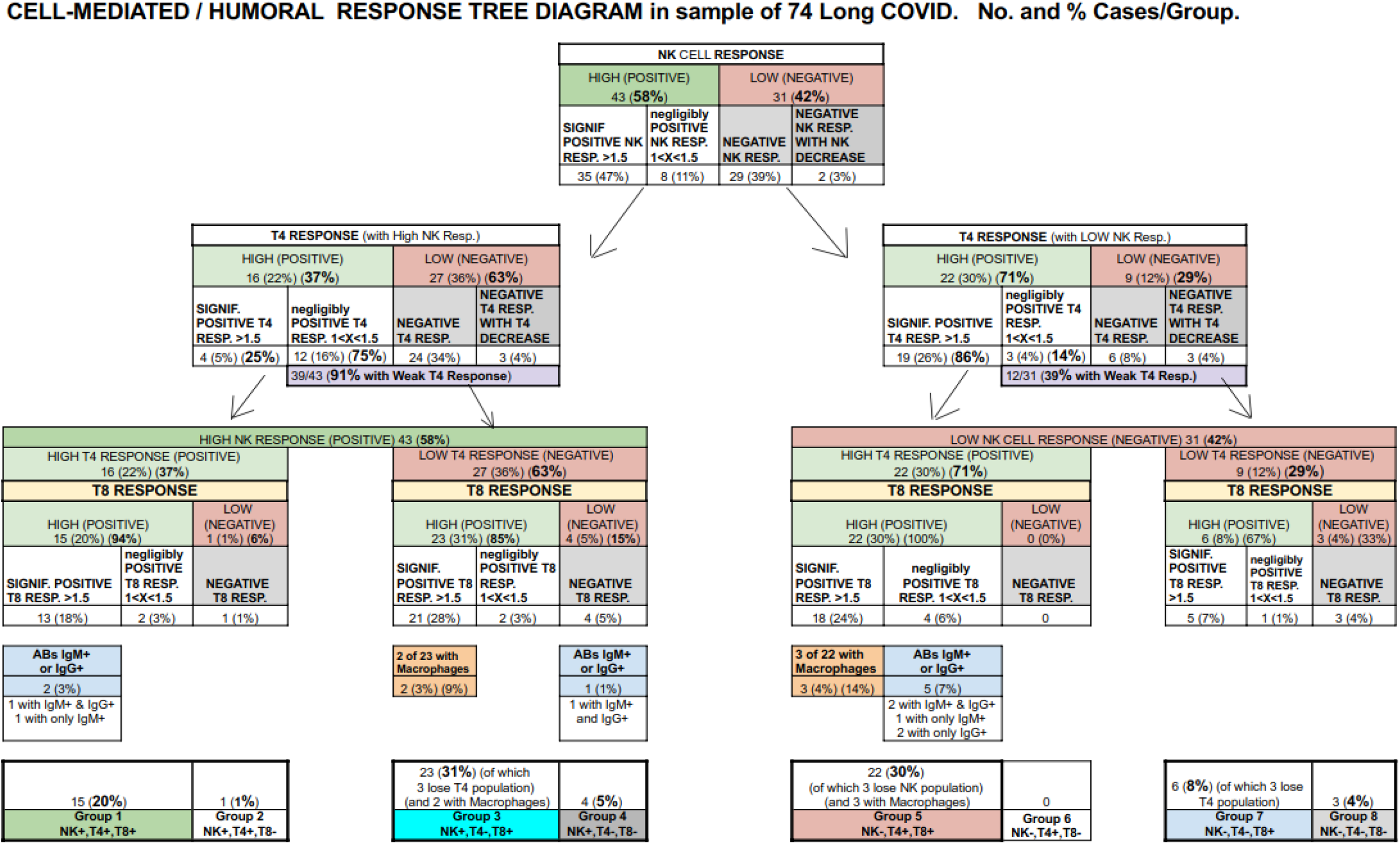
CLASSIFICATION (tree diagram) of GROUPS ACCORDING TO NK IMMUNE PROFILE → T4 → T8

The majority immune profile is that of **group 3** (NK+, T4-, T8+) with 23 patients (31% of 74), followed closely by **group 5** (NK-, T4+, T8+), with 22 participants (30%). The third-ranked group in terms of number of patients is **group 1** (NK+, T4+, T8+) with 15 patients (20%), and the rest of the groups are in the minority with 6 patients (8%) in **group 7** (NK-, T4-, T8+), 4 (5%) in **group 4** (NK+, T4-, T8-), 3 (4%) in **group 8** (NK-T4-, T8-), 1 (1%) in **group 2** (NK+, T4+, T8-) and 0 in **group 6** (NK-, T4+, T8-).

**Macrophage populations** were detected in the two largest groups (2 out of 23 cases: 9% in group 3, corresponding to 3% of all patients, and 3 out of 22 cases: 14% in group 5, 4% of total).

A **decrease in CD4+ T lymphocyte populations** when exposed to viral antigen was identified in group 3 (3 out of 23 cases: 13%) and group 7 (3 out of 6 cases: 50%). A decrease in NK populations was detected in group 5 (2 out of 22 cases: 9%).

The presence of a humoral response or **antibodies** is divided between **group 1** (2 out of 15 cases: 13%, one with IgM and IgG+ and another with only IgM+), **group 4** (1 out of 4 cases: 25%, with IgM and IgG+) and **group 5** (5 out of 22 cases: 23%, two with IgM and IgG+, one with only IgM+ and another with only IgG+), with no incidences in the largest group, group 3 (23).

As regards the use of **medication** recorded for 15 patients (20%), 8% of patients reported taking antihistamines, with 5% on Montelukast, 4% on oral corticosteroids, 3% on immunosuppressive treatment and 1% on antiviral treatment for Herpesvirus. None of the participants were receiving antiretroviral treatment for SARS-CoV-2, with only one case in the group where the patient did not have a specific immune response (group 8) and was being treated with Montelukast.

Table 4 shows the different diseases and symptoms on the survey in relation to their immune profile classification. Significant differences could not be established for the larger groups, although it would be interesting to be able to analyse these variables with a larger sample size.

**TABLE 4:**
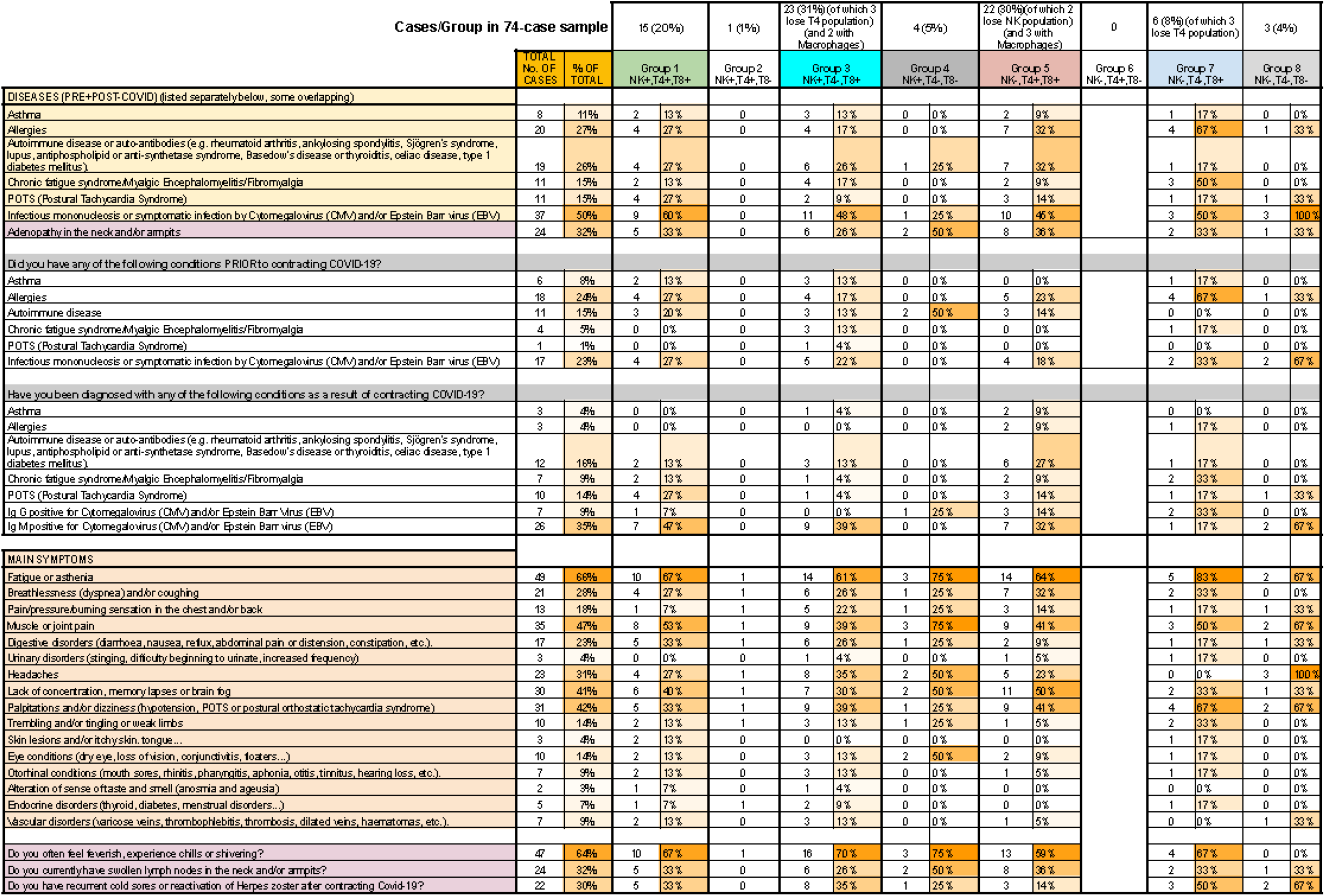
INCIDENCE BY GROUP OF CERTAIN DISEASES (PRIOR TO OR DIAGNOSED FOLLOWING COVID) WITH NO. OF CASES AND % OF MAIN SYMPTOMS

## DISCUSSION

In our sample of those affected by long COVID, 82% are still presenting symptoms more than 9 months (94.6% more than 7 months) after their initial COVID-19 episode. Patients are mainly middle-aged women, who display a higher percentage of illnesses related to “abnormal” immune responses (asthma, allergies and autoimmune disease), either preceding SARS-CoV-2 infection (43%) or following it in 23% of cases. This profile would match that of patients diagnosed with autoimmune diseases^(15)^ and it statistically corroborates the knowledge acquired about the possible aetiology of LC based on the persistence of the SARS-CoV-2 virus^(12)^.

### HUMORAL RESPONSE IN LC PATIENTS

This profile of altered immune response could contribute to the fact that a high percentage (81%) of patients with persistent symptoms previously present negative serology for SARS-CoV-2, despite being infected. This is a much higher rate than that found in the general population after recovering from COVID-19 (10%), as identified in our previous survey carried out with more than 200 infected individuals belonging to the Long COVID ACTS group (Long Covid Autonomous Communities Together Spain)^(12)^.

At the time of this analysis, humoral response is negative in 89% of cases, although it should be noted that a subgroup of patients (8%) are IgM-positive for SARS-CoV-2 after an average of 9.5 months from the start of symptoms, thus supporting the hypothesis of viral persistence. However, some of them had previously tested positive for antibodies and these are not detected in the current analysis; they have even lost them in less than 1 to 3 months.

Although on the whole a specific antibody response to SARS-CoV-2 is not generated, the possible persistence of a viral reservoir would prolong immune system stimulation, leading to the maintenance of permanently activated lymphocytes (not necessarily those specific to SARS-CoV-2), and this would make them susceptible to autoreactivity, inducing a response to internal body structures and autoimmune phenomena. This would align with the fact that with many persistent viral infections there is a delayed response in the production of neutralising antibodies, and they are also characterised by producing a high level of non-specific antibodies^(16)^.

A recently published article^(17)^ finds that patients with long COVID have functional antibodies against receptors linked to G protein, also found in patients with Chronic Fatigue Syndrome or POTS/dysautonomia, with both diseases diagnosed in LC patients after acute SARS-CoV-2 infection, and picked up in our sample as a condition that pre-dated or followed the infection. These are also illnesses of uncertain aetiology, sometimes triggered by a viral infection, yet they do have traits that differentiate them from LC ^(12)^.

This article^(17)^ explains that a combination of antibodies and ischemic cofactors or inflammations can work to maintain an inflammation process, which explains the persistence of symptoms in LC patients, indicating and indeed demonstrating that certain antibodies can affect the maturing and degranulation of cardiac **mast cells**, contributing to said inflammation.

By some mechanism or other, this lack of immune system response regulation could itself be linked with some patients’ allergy profile, as seen in 27% of the sample, or it could be a predisposing factor as 24% presented it as underlying. Likewise, SARS-CoV-2 infection could trigger mast cell activation syndrome^(18)^ in them, linked to acute and chronic COVID-19 in the literature, with a clinical picture which at times overlaps with that of long COVID^(18)^.

### IMMUNE CELL TYPING IN LC PATIENTS

This possible change to the immune system identified in LC patients, together with scarce antibody production, leads us to analyse lymphocyte typing and cellular response.

Altered immune cell typing can be identified in many of the patients, with high levels of CD3 lymphocytes found in more than one third of cases, and high CD8 levels in a quarter of them. However, B lymphocytes and monocytes are low in almost a third of our sample and high in a small percentage (15%), although these two cell populations were not differentiated by the laboratory, and it would be interesting to be able to do so in future studies.

Another important point is that a third of LC patients surveyed presented reduced values of T CD4 lymphocytes, a basic pillar of the specific immune response, and almost two thirds showed an increased NK population. None of the patients were found to have high lymphocytes T4 nor low NK values.

This increased level of NK cells infers a persistent inflammatory state mediated by innate cell immunity in a significant number of patients with LC, which could cause many of the symptoms reported. The trigger that could provoke this permanent immune response could be the persistent presence of the virus in reservoirs.

In addition, it is interesting to note that in a minor percentage of participants, macrophage populations were detected in peripheral blood when their normal location is in body tissues.

This finding could indicate the possible involvement of macrophages in the illness as something requiring further research. They are likely related to viral persistence, with possible reservoirs in body tissues, where their precursors, monocytes^(19,20)^ are transformed, as has already been referred in the case of feline Coronavirus^(21)^.

It has been described that chronic inflammation and immune response in persistent viral infections is associated with several immune dysfunctions including aberrant activation of T lymphocytes, senescence and cell exhaustion, dysfunctional B lymphocyte response, polyclonal B lymphocyte activation, changes to innate immunity and lymphoid architecture disruption^(16)^.

### SPECIFIC CELLULAR RESPONSE TO SARS-CoV-2 IN LC PATIENTS

Despite 76% of those surveyed not previously testing positive for SARS-CoV-2 by means of a microbiological or serological test, 96% showed specific cellular immunity to SARS-CoV-2 an average of 9.5 months after infection: 89% of this response corresponds to T8 lymphocytes and 58% to NK cells. The T4 lymphocyte response, however, is lower, and negative in 49% of cases.

Furthermore, given that no significant differences were detected regarding typing and kind of specific cellular response between the group that had previously tested positive through a diagnostic test and those who had not, this proves that specific cellular immunity to SARS-CoV-2 can be a good tool, in the vast majority of cases, to confirm infection (COVID-19) up to over nine months since the initial episode in those people who present with LC symptoms and negative microbiological tests, given that this indicates there has been contact with the virus despite the absence of an antibody titre.

The presence of cellular immunity could be the reason why such low numbers of LC patients present with reinfection and, in the small number of those who go on to test positive in a nasopharyngeal PCR after their first infection, it has not been proven if this is due to reinfection or increase in transitory viral load in a persistent infection.

A study of 188 people (80 men and 108 women) ranging from being asymptomatic to mild, moderate and severe cases of COVID-19, found that T4 and T8 lymphocytes specific to SARS-CoV-2 had a half-life of 3 to 5 months^(22)^. Another study concludes that changes in B and T cell function following hospitalisation due to COVID-19 could affect immunity in the longer term (up to 6 months) and contribute to some persistent symptoms observed in patients convalescing from COVID-19.^(23)^

Our findings are that 90% (67 out of 74) of patients with LC still have specific cellular immunity more than 7 months after primary infection (78% after more than 9), and there is scope to consider that in the majority of LC patients, cellular immunity could go on for a longer period. The fact that there is prolonged specific T cell activity could be an indirect indicator of viral persistence in LC patients.^(23)^

Half of the surveyed LC patients appear to have a ‘defective’ CD4+ T lymphocyte response, which would prevent CD8+ T lymphocytes from activating adequately, thereby affecting proper antibody production. To compensate, NK cells would increase to try to fight the virus through innate response. It has been described that the persistence of another virus results in CD4+ T lymphocytes being altered or redirected^(16)^.

More than two thirds of LC patients surveyed had a weak (20%) or non-existent (49%) CD4+ T lymphocyte response, this being the predominant cytotoxic response mediated by CD8+ with a specific response in 89% of cases, significant in the vast majority of them (77%).

These results totally contradict those reported in the literature, in which the CD4+ T lymphocyte response to SARS-CoV-2 in convalescing patients is more prominent than that of T8 lymphocytes ^(24,25)^ and has been associated with primary control of the infection^(26)^.

Consequently, in response to SARS-CoV-2, fewer CD8+ T lymphocytes can be found circulating compared with T4 lymphocytes^(24-26)^. The coordinated response of CD4+ T and specific CD8+ T lymphocytes and antibodies protects against SARS-CoV-2, but an uncoordinated response frequently fails to control the infection ^(25)^.

As the symptoms of those with reduced CD4+ T lymphocytes and the rest of the LC patients is identical, and increased NK levels are frequently found in both cases (without a statistically significant difference), it is probable that patients with a normal range of CD4+ T lymphocytes could present other defects in their activation mechanism.

This would be supported by the fact that, including those who present a normal number of CD4+ T lymphocytes, the vast majority present a specific cellular response to SARS-CoV-2 of less CD4+ T lymphocytes than CD8+ T lymphocytes, contrary to what is described in the literature.

It has been described that in chronic infection of Lymphocytic choriomeningitis virus (LCMV), specific T cells enter an attenuated (exhausted) state characterised by a loss of their ability to proliferate, produce key antivirals, produce stimulating cytokines and a loss of cytolytic activity^(16)^. In LCMV a direct link has been found between IFN-I signaling, immune suppression and viral persistence. IL-10 is one of the immune system’s key negative regulators during multiple persistent viral infections. It would be interesting to study its levels in LC patients^(16)^.

There is an additional phenomenon observable in some of the LC patients studied who have a negative CD4+ T or NK response, where a reduction in CD4+ T and NK lymphocyte populations when exposed to viral antigen (8% for CD4+ T and 3% for NK) is noted.

This reduction in CD4+ T lymphocytes in response to re-exposure to the viral antigens could be explained as a mechanism known as AICD (Activation-Induced Cell Death)^(27)^, in which T lymphocytes, activated by a constant antigen stimulus, become more susceptible to death by apoptosis, and are then eliminated once the infection has been fought, thereby recovering immune system homoeostasis. This death of T lymphocytes effectors by apoptosis is produced mainly after continued exposure to the antigen responsible for producing their initial activation. When it comes to NK, these have been subject to much less research and definition, but it appears they could also suffer from AICD. Our sample did not detect any cases of this happening to CD8+ T lymphocytes.

With reference to COVID-19, we found an increased secretion of some cytokines, which correlates with the reduction of T lymphocytes detected in patients infected with SARS-CoV-2 in the acute phase, but which might also play a pathological role in chronic inflammation and infection, like IL-6. Other cytokines include TNF-α which causes apoptosis in T cells, and IL-10, which can induce exhaustion in these cells, that has found to be preventable in animal studies of chronic infection, where this cytokine was blocked^(28)^.

It would be interesting to study the levels of these cytokines in LC patients, and also to research the effect of the virus on lymphocyte populations in LC, especially CD4+ T lymphocytes, reduced in one third of our sample, and also to assess if these are susceptible to SARS-CoV-2 infection, as has been previously described in the case of SARS-CoV-1^(29)^.

This immunological alteration which exists in LC patients could be the cause of the reactivation of other latent viruses such as Herpes, CMV or EBV (previously researched by this group^(12)^), confirmed statistically in our survey. The loss of T lymphocyte functionality in persistent viral infections consists of the immune system’s reduced ability to mount a de novo immune response against co-infectious pathogens, resulting in the host having reduced capacity to fight infections and potentially to form new memory T cells^(16)^.

It is notable that 64% of those surveyed currently present with frequent fever, chills and shivering, and almost a third report Herpesvirus (zoster or simplex type 1) reactivation or adenopathy. Moreover, half present previous symptomatic processes or a current diagnosis indicated by serology (mostly IgG) relating to the presence of latent viruses such as CMV or EBV.

It would be interesting to study possible reactivation of latent viruses in LC patients that pre-date SARS-CoV-2 infection using PCR, as the majority present abnormal serological response and it is therefore highly likely they do not conform to the ‘standard’ serological interpretation of virus reactivation (presence of IgM).

Viral persistence could be the trigger for the immunological alteration observed in the specific cellular response to SARS-CoV-2 in LC patients, with this state of high inflammatory response causing the majority of clinical presentations.

If we analyse specific immune response profiles to SARS-CoV-2 with the **classification** that has been established, most cases are in groups 3 (NK+, T4-, T8+) and 5 (NK-, T4+, T8+) accounting for 31% and 30% of participants respectively. Both share positive T8 response, but T4 and opposite NK responses which are probably compensatory.

In group 3 (NK+, T4-, T8+), the absence of specific CD4+ T lymphocytes for SARS-CoV-2 suggests that the specific CD8+ T generated did so without their help, and therefore are probably unable to carry out their function as well as if there had been good cooperation between both lymphocyte populations. For this reason, in this group it is possible that NK cells persist to compensate for the lack of correctly functioning CD8+ T. It should be noted that CD8+ T and NK, despite being activated differently, both then perform the same function: they eliminate cells infected by the virus. In this group none of the patients presented a humoral response, but it was the majority immune response profile (3 out of 7: 43%) in patients who had a positive antibody titre in a previous diagnostic test and have lost them at the time of current analysis, after a mean of 5 months.

Group 5 (NK-, T4+, T8+) did resemble a normal immune response. If, in response to the virus, CD4+ T cells activate and can collaborate correctly with CD8+ T cells, the innate (NK) response reduces because it is no longer necessary. However, in this case CD4+ T do not appear to be activating correctly in terms of their collaborative function with B lymphocytes, given that antibodies can only be detected in 23% of patients (5 out of 22).

It would be interesting to be able to study T4, T8 and NK phenotypes (effectors, memory, adaptive, exhausted…).

Group 1 (NK+, T4+, T8+), with its specific response by the 3 lymphocyte populations, is the third most frequent group (20%), and 2 out of its 8 patients have positive antibody titres.

Group 8 (NK-, T4-, T8-) comprises 3 patients (4% of the total) whose immune response, both cellular and humoral, was negative at the time of carrying out research analysis, and all of them presented symptoms for over 9 months. However, we can confirm that not having a specific immune response at present does not rule out illness given that one of them was previously diagnosed with Covid-19 through PCR and serology.

It is also worth noting that all three have processes associated with viral reactivation, either because of a history of mononucleosis symptoms or the presence of IgG for CMV/EBV, and one of them currently presents with adenopathy and recurrence of Herpesvirus simplex type 1 or zoster. Reactivation of these conditions as a result of SARS-CoV-2 infection, with no detectable specific immune response at present, could also contribute to these persistent symptoms. Immunomodulating medicines do not appear to be a contributory factor, as out of 4 being treated with Montelukast, only a single case belongs to the group where this response is negative.

The possible abnormal cellular and humoral immune response to the virus detected in LC patients could be the source of why the patients are unable to effectively eliminate the virus. It may remain in reservoirs, causing a state of permanent inflammation related to the innate immune response, or the viral persistence itself may cause an altered immune response.

This study has **limitations and strengths**. Its main strength is that it is an innovative study which aims to start/launch a research hypothesis for LC. However, there are also limitations: the principal limitation being the lack of a control group matched by age, sex and time passed since overcoming COVID-19, without presenting with persistent symptoms. Other limitations presented by this study are that the data analysed were gathered with a survey and the values of B lymphocytes and monocytes have not been provided by the laboratory in an independent manner. In addition, it is useful to note that the cellular response was recorded as being negative, positive and negligible (1-1.5%) or positive and significant (>1.5%) and it would be useful to quantify the value of IFN-gamma production by NK cells in response to SARS-CoV-2 compared to the NK cell baseline, this being a more non-specific innate response. This occurs only in group 4 which has 4 patients, and one of them has a specific antibody titre for SARS-CoV-2.

## CONCLUSION

To conclude, it could be argued that the immune system appears to have an important involvement in the development of long COVID, since patients display a higher percentage of illnesses related to “abnormal” immune responses and most of them had negative serology for Covid-19. Despite the high percentage without previously testing positive for SARS-CoV-2, most of them showed specific cellular immunity to SARS-CoV-2 more than 9 months after infection, this being the predominant cytotoxic response mediated by CD8+, with a weaker or non-existent CD4+ T lymphocyte response, results that contradict those reported in the literature in convalescing patients associated with primary control of the infection. This fact probably leads to an increased innate (NK) response to compensate the lack of immune cells cooperation, what infers a persistent inflammatory state, which could cause many of the symptoms reported. Altered immune cell typing count had been identified in many of the patients as well with detection of macrophage populations in the peripheral blood in a minor percentage of participants. We defend the hypothesis of viral persistence as a possible cause or consequence of all this alteration of the immune system, but it would be necessary to expand the study with a control group.

We found two predominant specific cellular immune response profiles to SARS-CoV-2, and it would be interesting to conduct a separate data analysis of monocytes and B lymphocytes, including a study of the presence of potential reservoirs of SARS-CoV-2 in monocytes and macrophages, and analyse subpopulations of lymphocytes and phenotypes of NK, CD4+ T and CD8+ T (effector, memory, adaptive, exhausted cells…). An autoimmunity study would also be useful for developing a better understanding of the disease’s aetiopathogenic mechanisms to enable effective treatments to be found.

## Data Availability

The authors assert that all procedures contributing to this work comply with the ethical standards of the Helsinki Declaration of 1975, as revised in 2008. All data were collected anonymously.

## Ethics statement

The authors assert that all procedures contributing to this work comply with the ethical standards of the Helsinki Declaration of 1975, as revised in 2008. Protocol was approved by the Clinical Research Ethics Committee of Institute of Research of Aragon, IIS-Aragon (protocol code PI21/278). All data were collected anonymously.

## Author contributions

NM, SG and ED were responsible for the conception, data collection, and design of the study. NM and BOB were responsible for data analysis. ED, SG contributed to the interpretation of data. NM and SG wrote the article, which was critically revised by all the other authors. All authors have approved the final version of the manuscript.

## Acknowledgments

We wish to thank Luis Martínez Lostao (immunologist), Alberto Duarte, as well as Sonia Bilbao, Mercedes Sánchez and Gemma Pereira, Teletest Lab Analisis and the LongCOVID ACTS (Autonomous Communities Together Spain) alliance for their participation. We wish also to thank the Primary Care Prevention and Health Promotion Network (RedIAPP-Health Institute Carlos III, Spain); Research Group B21_R17 of the Department of Research, Innovation and University of the Government of Aragon (Spain); Feder Funds “A way to make Europe”, for their support in the development of the study

